# Data Mining Approach to Analyze Covid19 Dataset of Brazilian Patients

**DOI:** 10.1101/2020.08.13.20174508

**Authors:** Josimar Chire

## Abstract

The pandemic originated by coronavirus(covid19), name coined by World Health Organization during the first month in 2020. Actually, almost all the countries presented covid19 positive cases and governments are choosing different health policies to stop the infection and many research groups are working on patients data to understand the virus, at the same time scientists are looking for a vacuum to enhance imnulogy system to tack covid19 virus. One of top countries with more infections is Brazil, until August 11 had a total of 3,112,393 cases. Research Foundation of Sao Paulo State(Fapesp) released a dataset, it was an innovative in collaboration with hospitals(Einstein, Sirio-Libanes), laboratory(Fleury) and Sao Paulo University to foster reseach on this trend topic. The present paper presents an exploratory analysis of the datasets, using a Data Mining Approach, and some inconsistencies are found, i.e. NaN values, null references values for analytes, outliers on results of analytes, encoding issues. The results were cleaned datasets for future studies, but at least a 20% of data were discarded because of non numerical, null values and numbers out of reference range.

## 1 Introduction

The outbreak of Coronavirus(Covid19) started with first cases on December 2019, in Wuhan(China). The first reported case[1] in South America was in Brazil on 26 February 2020, in São Paulo city. The strategy to stop the infections in the country was a partial lockdown to avoid the propagation of the virus.

On 28 January 2020, Ministry of Health of Brazil reported a suspected case of Covid19 in Belo Horizonte, Minas Gerais state, recently one student returned from China [2], [3]. The same day were reported two suspected cases in Porto Alegre and Curitiba [4]. The first confirmed COVID-19 case [5] were reported in Brazil, a man of 61-year-old who returned from Italy. The patient was tested in Israelita Einstein Hospital in Sao Paulo state. On 14 May[6], more than 200 000 cases were confirmmed, this number double during the first days of May.

Until August 11, the numbers of Brazil^1^ are: total of 3,112,393 cases, with an increasing rate of new cases of 44,255(+1.4%) and a total of 2,243,124 recovered cases.

Nowdays, many scientists are working around coronavirus covid19, but searching for conducted studies in South America, there is only a few number. After a searching in IEEX Xplorer using coronavirus, covid19 terms, one paper with Brazilian Affiliation is found [7], related to data augmentation for covid19 detection. Considering a preprint repository related to Medicine(Medxriv), using terms: covid19, coronavirus, data mining more than 50 papers are found.

The table 1 presents the top 10 results of MedxRiv query. Four of this papers is a conducted study for South America countries and there is any work analyzing Brazilian context. In spite of, there is 4 papers with Brazilian Affiliation.

**Table 1.**
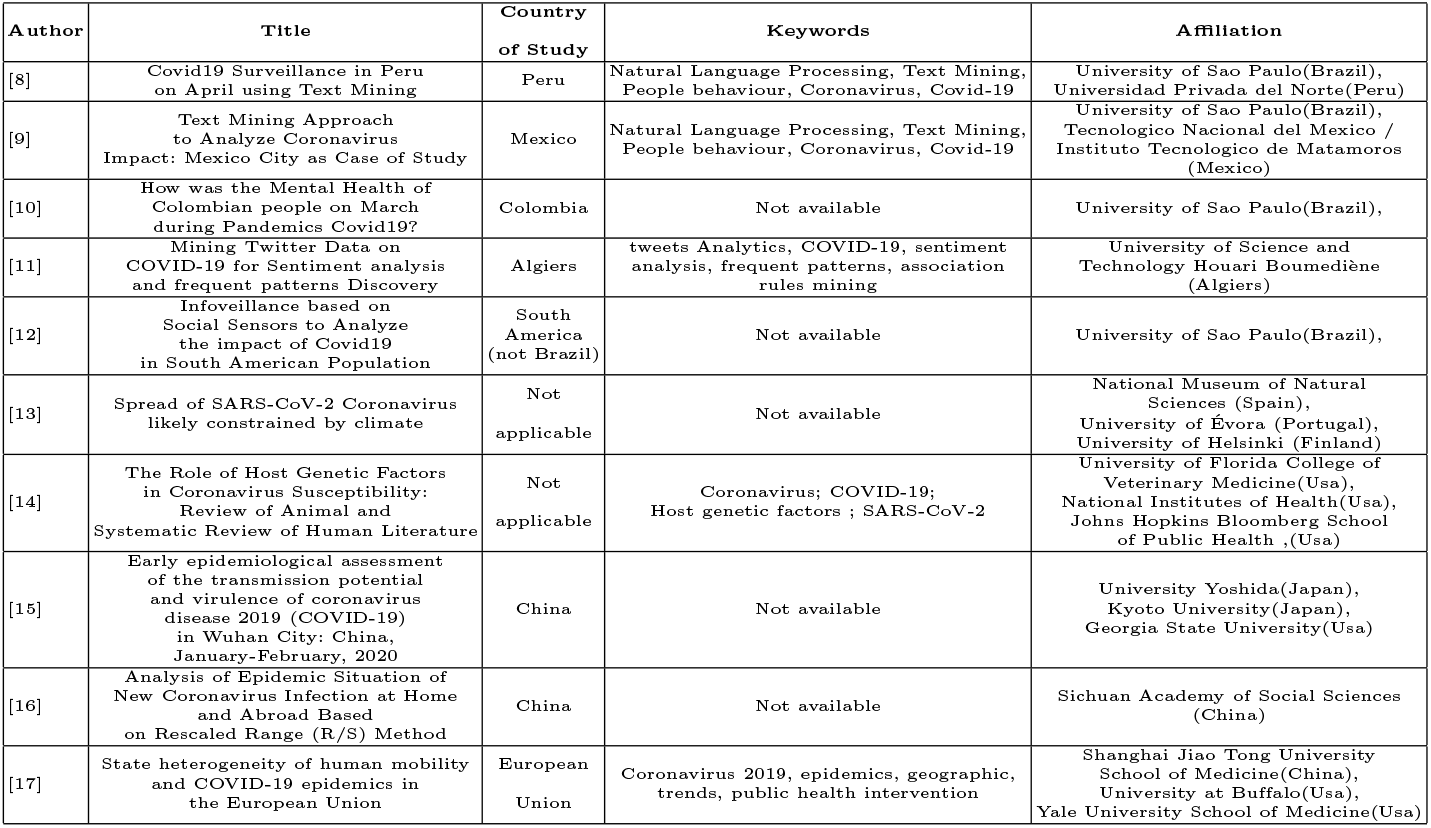
Ten results of Medrxiv Query about covid19 papers in South America

Considering, the previous evidence it is necessary to conduct studies with Brazilian data, then the initiative of Fapesp is valuable to foster research on covid19 topic. The actual paper uses Data Mining Approach to perform an exploratory analysis of the dataset of Brazilian patients of Sao Paulo State. The methodology to explore data is presented in Section 2, the experiments and results in Section 3. Conclusion states in Section 4, final recommendations and future work are presenten in Section 5, 6.

## 2 Methodology

The conducted work follows a methodology inspired in CRISP-DM[18]. The image 1 presents the flow between the phases of the exploration.

**Fig. 1.**
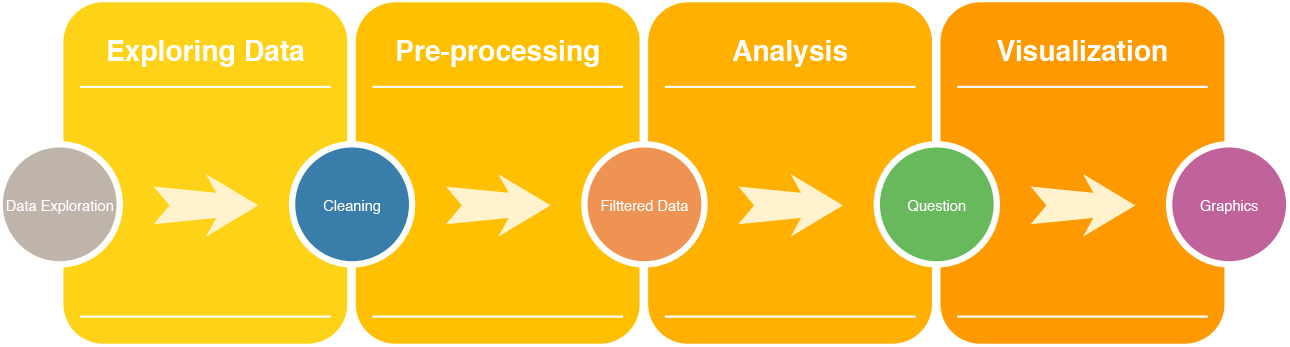
Methodology

### 2.1 Exploring data

This step involves: check format files, open the files using a Language Programming or a tool. Review number of registers or rows per each file. Check existence of null values, check kind of each variable or field. For this step, Python Language Programming and pandas package are used to manipulate the data.

### 2.2 Pre-processing Data

This step is related how to deal with data before of generate graphics for analysis.

– If a specific variable must be numerical, but there is string values, so it is discarded
– If null values are found, a discarding process must be considered.
– If range reference for one exam, analytes is null then the analysis is not possible.

### 2.3 Analysis

Using clean data is possible to answer some questions related to age distribution, sex distribution, distribution of results to detect anomalies or outliers. The questions can require a kind of specific graphic to suppot analysis.

### 2.4 Visualization

Considering distribution of few classes, a pie chart is useful to check proportions, subsection 3.3, 3.8. For age distribution, bar plot can show how is the distribution, see subsection 3.4, 3.5, 3.6. The analysis is dozen of values can be supported for boxplot graphics, in subsection 3.9, 3.10.

## 3 Experiments and Results

### 3.1 Datasets

The release of the datasets is the result of collaboration between Research Foundation (FAPESP)[19], Fleury Institute, Israelita Albert Einstein Hospital, Sirio-Libanes Hospital and the University of Sao Paulo. The goal is to contribute and promote research related to Covid19. The datasets share the data dictionaries of Patients(see Tab. 1), Test (Tab. 2).

**Table 2.**
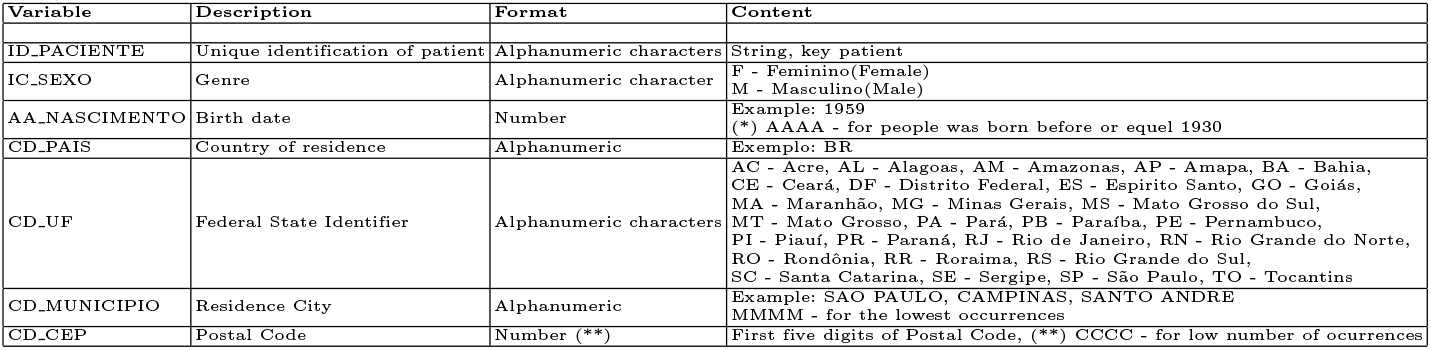
Data Dictionary of Patient Dataset-Einstein, Fleury, Sirio-Libanes Hospital

**Table 3.** Data Dictionary of Tests - Einstein, Fleury, Sirio-Libanes Hospital

| Variable Name | Description | Format | Content |
| --- | --- | --- | --- |
| ID_PACIENTE | Unique identification of patient | Alphanumeric character | String, patient key |
| DT_COLETA | Exam collection date | Date (yyyy/MM/dd) | Date |
| DE_ORIGEM | Origin of patient | Alphanumeric character (4) | HOSP - Exam made in a hospital |
| DE_EXAME | Description of Exam | Alphanumeric | Example: HEMOGRAMA(blood count) |
| DE_ANALITO | Analyte description | Alphanumeric | Example: Eritrócitos(Erythrocytes), Leucócitos(Leukocytes), Glicose(Glucose) |
| DE_RESULTADO | Result of exam, related to DE_ANALITO | Alphanumeric | If DE_ANALITO requires numerical values, Integer ou Float<br>If DE_ANALITO requeries qualitative, String with restrict domain |
| CD_UNIDADE | Unit of measurement | Alphanumeric | String<br>Exemplo: g/dL (grams por deciliter) |
| DE_VALOR_REFERENCIA | Reference values for DE_RESULTADO | Alphanumeric | String - Reference value for de_analito in the population *MinValue*, *MaxValue*<br>Não Detectado(Not detected)/Detectado(Detected)<br>Example for glucose: 75 to 99<br>Example for progesterone: until 89 |

The size of dataset are presented in Table 3 for three data sources. SL Hospital provided a dataset about outcomes of the patients.

**Table 4.** Features of Dataset

|  | Einstein Hospital | Fleury | SL Hospital |
| --- | --- | --- | --- |
| **Patient(size)** | 43,562 | 129,596 | 2,731 |
| **Test(size)** | 1,853,695 | 2,496,591 | 371,357 |
| **Test(Dates)** | 2020-01-01<br>to 2020-06-24 | 2019-11-01<br>to 2020-06-15 | 2020-02-26<br>to 2020-06-27 |
| **Outcome(size)** | - | - | 9,634 |
| **Outcome(Dates)** | - | - | 2020-02-26<br>to 2020-06-29 |

### 3.2 Exploration

This subsection present some graphics to describe data and let posterior analysis, besides the requeriment of some graphics related to distribution, i.e. bar plot, boxplot.

#### Description of datasets

The Figure 2 is presented with counting values, unique values, top for each field. The name of columns were transformed to lowercase to have an uniform name of fields.

**Fig. 2.**
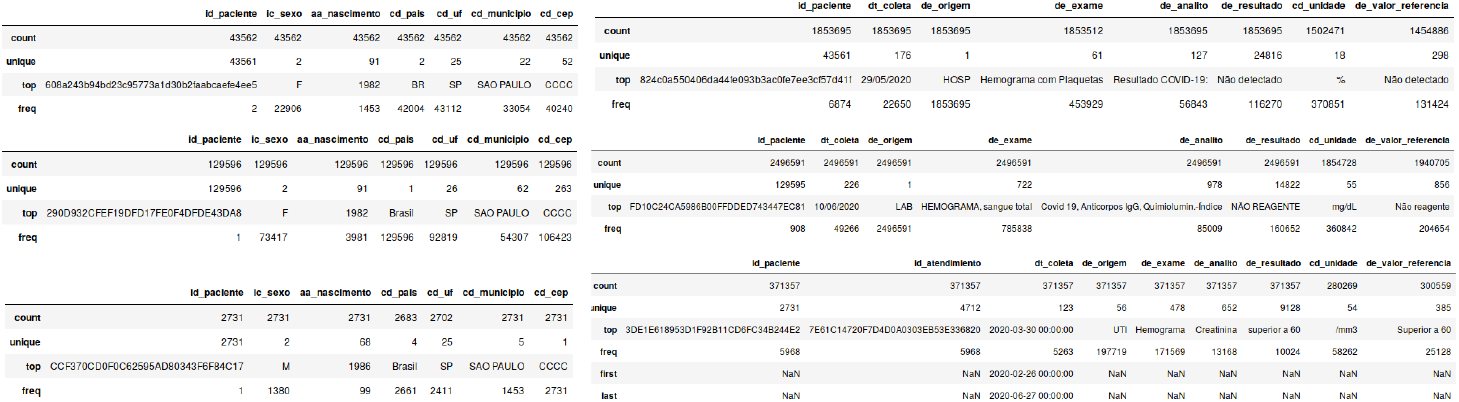
(a) Einstein, (b) Fleury and (c) SL Datasets Description

– Figure 3.b presents a different number of id_paciente in patient dataset and exam dataset, 129596(patient) 129595(exam).
– Einstein and SL Hospitals(cd_pais) presents people living in countries different than Brazil.
– The most frequent age of patients is: 38(Einstein, Fleury) and 34(SL).
– Female patients are higher in number in Einstein, Fleury.
– Most frequent cd_uf, cd_municipio is Sao Paulo State or city and CCCC is most common in Postal Code, so this places do not have meaningful number of ocurrences.
– Einstein and Fleury have a unique de_origem: Hosp, Lab respectively. But SL Hospital has 56 different.
– The exam hemograma(blood count) is the most frequent in the datasets, and de_analito more frequent in Eistein, Fleury are related to **Covid19**.
– Eistein has the lowest number of different de_exame(61), de_analito(127). Fleury has the highest de_exame(722), de_analito(978). SL has de_exame(478), de_analito(652). Therefore, numer of de_valor_referencia are related.
– SL Hospital presentes NaN(Not a number) values, then it is possible find NaN values in the datasets.

### 3.3 Sex Distribution

Female population is slightly bigger than male population in Einstein, Fleury but SL presents male population bigger for 0.05%(29 people), see Fig. 3.

**Fig. 3.**
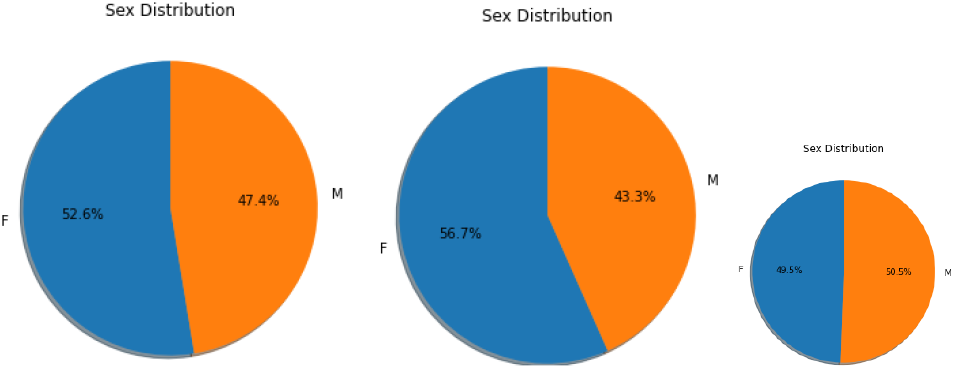
Sex Distribution(Einstein, Fleury and HL)

### 3.4 Age Distribution

Datasets of Einstein, Fleury have younger patients from 0 to 14 until 89 but SL Hospital only from 14 to older(86), this graphics are presented in Fig. 4

**Fig. 4.**
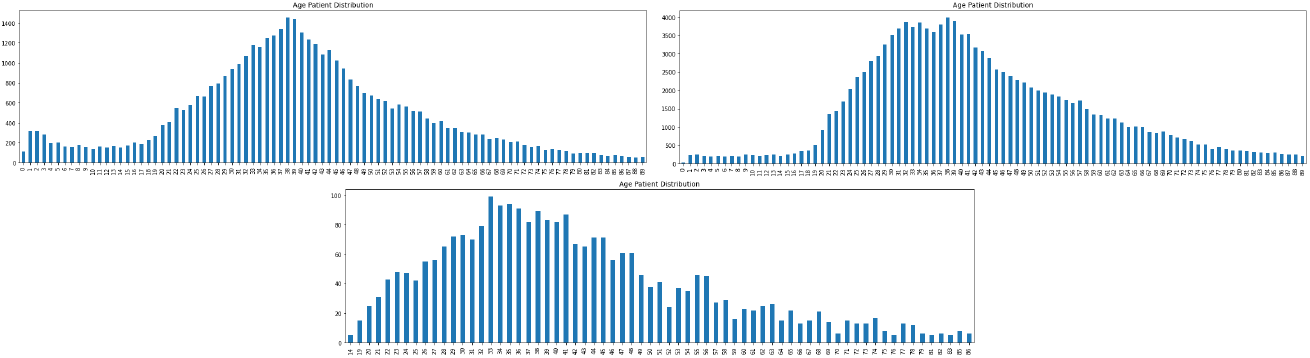
Age Distribution (Einstein, Fleury, SL)

### 3.5 Date Collection of Exams

The graphic Fig. 5 presents the number of collect exams per day and month, Einstein presents an increasing number from January to June, Flury a decreasing from January to April but a peak on May, June. Besides, SL Hospital has an increasing from February to June.

**Fig. 5.**
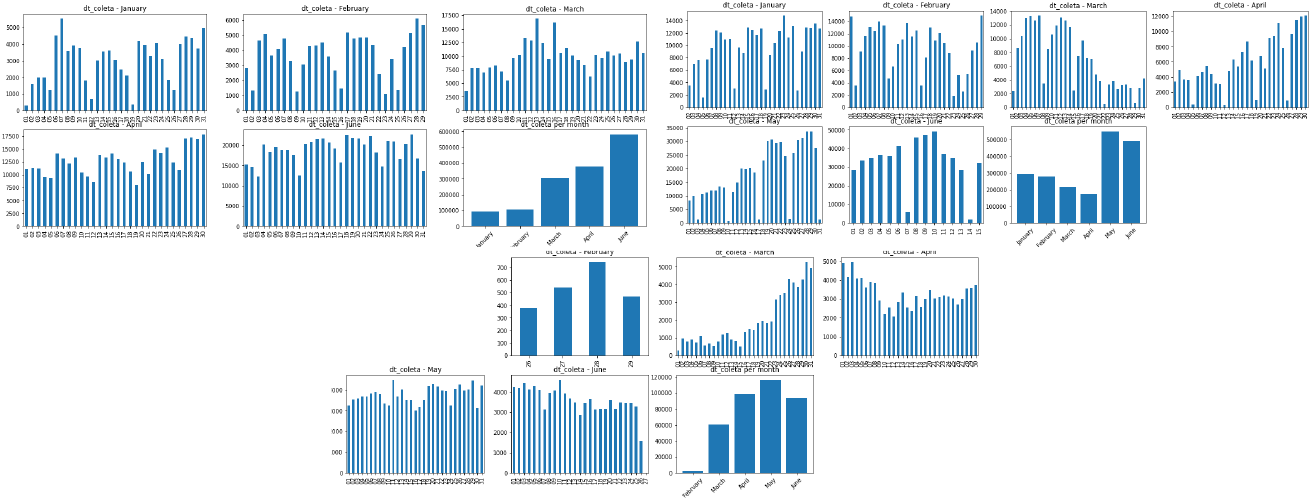
Date Distribution (Einstein, Fleury, SL)

**Fig. 6.**
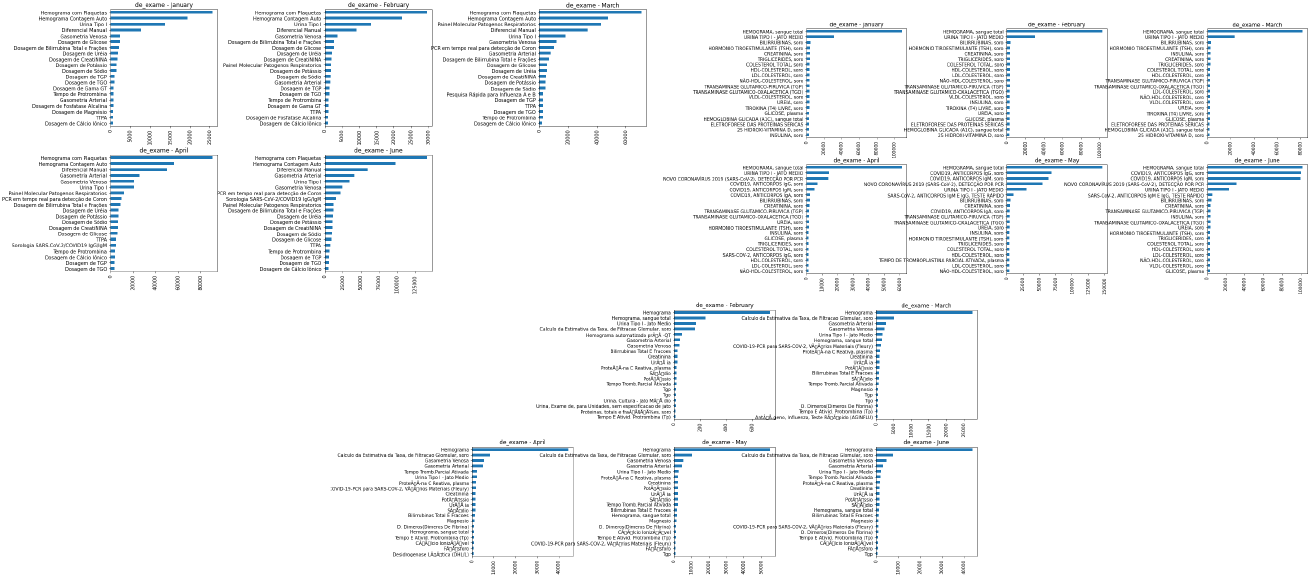
Exam Distribution (Einstein, Fleury, SL)

### 3.6 Most frequent exams per month

To answer what were the most frequent exams during the month of each dataset, graphic Fig. 7 presents the 20 most frequents.

– Three datasets has blood count exam on the top of each month.
– Only Fleury has exams related to covid19 detection on April, May, June on the top 5.
– There are many kind of exams related to covid19 for Hospital, i.e. PCR, Sorologia SARS-Cov-2/Covid19 (Einstein). Fleury has NOVO Coronavirus 2019, Covid19 Anticorpos lgG, lgM, lgA and more. SL Hospital has Covid-19 PCR para Sars-Cov2 and a problem with encoding is detected in this dataset.
– For the previous reason, each dataset is studied separately.

**Fig. 7.**
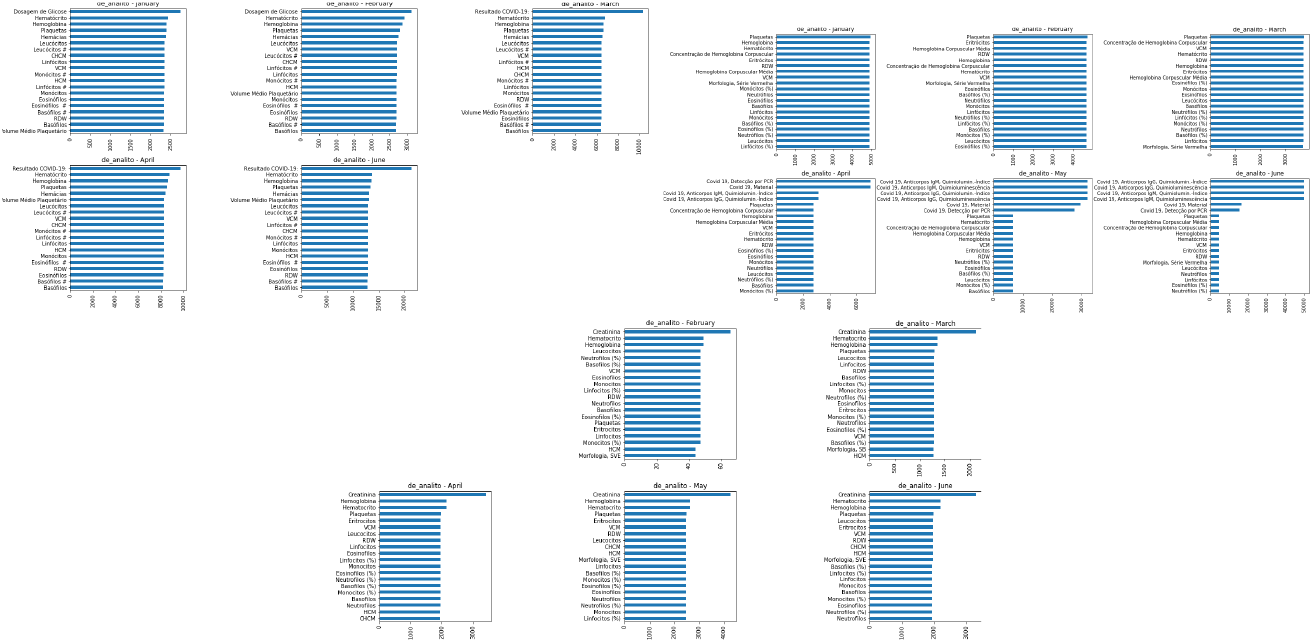
Analyte per month(Einstein, Fleury, SL)

### 3.7 Most frequent analyte per month

Einstein and Fleury presents analytes related to covid19, i.e. resultado covid19, Covid19 deteccao por PCR, Covid19 material and more. Again, Fleury presents a variety of names for analytes related to covid19. And SL Hospital does not have any in the top 20.

### 3.8 Covid19 Analytes Distribution

Considering analytes related to covid19, graphic 8 presents the number of detected/not detected during the months for Hospital Einstein. Fleury and SL do not have an standardized outputs of covid19 exams, therefore is not possible to generate the graphics yet.

**Fig. 8.**
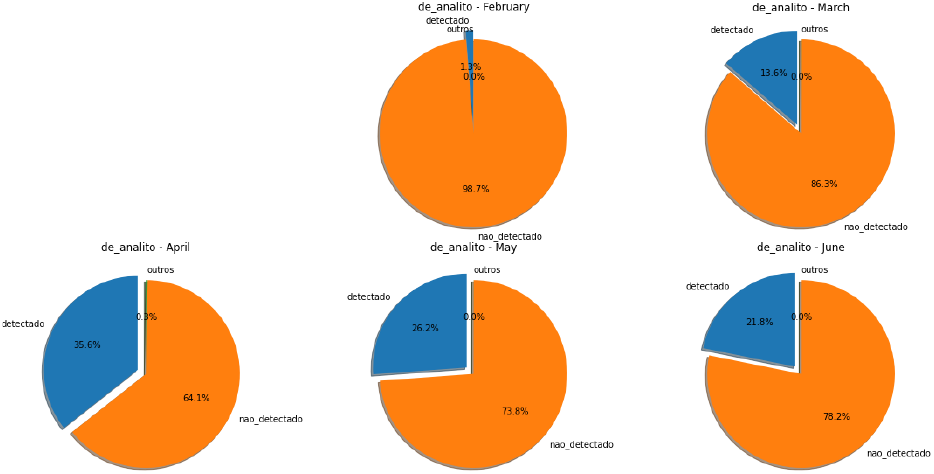
Analyte per month(Einstein)

### 3.9 Boxplot of most frequent exams

Considering top 14 of de_analito and de_resultado, the graphic Fig. 9 is presenting boxplot of the values of Einstein Hospital. It is necessary not to consider qualitative values, then only numerical values were used to build the plot. Analyzing the graphic is remarkable to many outliers in many of analytes, then a cleaning process is necessary.

**Fig. 9.**
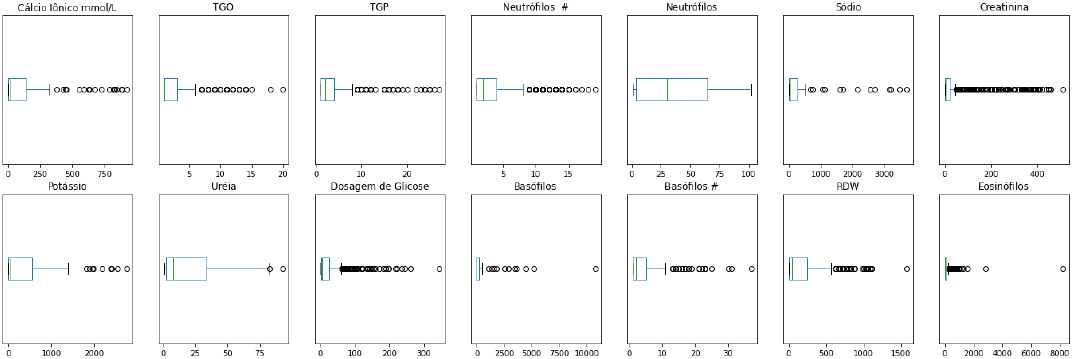
Boxplot of top 14 analytes (Einstein)

Splitting data of covid19 detected and no detected, figure Fig. 10 is presented. Again, outliers are present in Fleury dataset. Red ones(detected), blue(not detected).

**Fig. 10.**
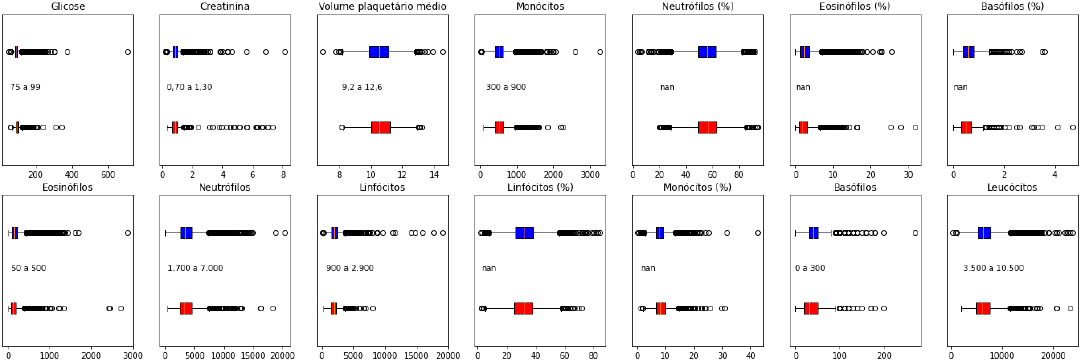
Boxplot of top 14 analytes(Fleury)

Using a cleaning process using standard deviation(std) is proposed, because the outliers are further than median and in normal case two or three times higher is considered an abnormal value but in this situation, to have a better visualization of boxplot was used 0.5*std(see Fig. 11) and 0.2*std(see Fig. 11) on Einstein dataset considering analytes with abnormal values.

**Fig. 11.**
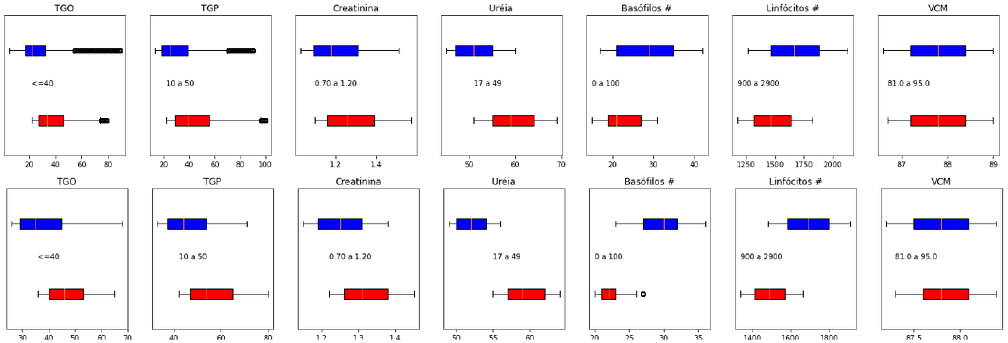
Boxplot of Cleaned dataset of Analytes with Abnormal Values, 0.5*std

### 3.10 Cleaning using Reference Values

The next graphics are created splitting Einstein dataset for genre. There is presence of NaN values in the reference value then these analytes are discared for the graphic, table 3.10 presents the no valid de_analito, it is a total of 8.

**Table 5.**
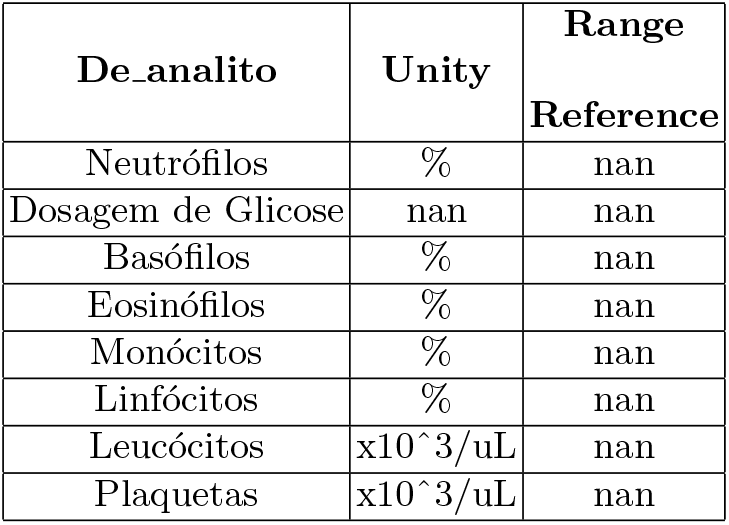
No valid de_analito for no valid reference range

Plotting the distribution(Fig. 12) for 30 most frequents analytes for men.

**Fig. 12.**
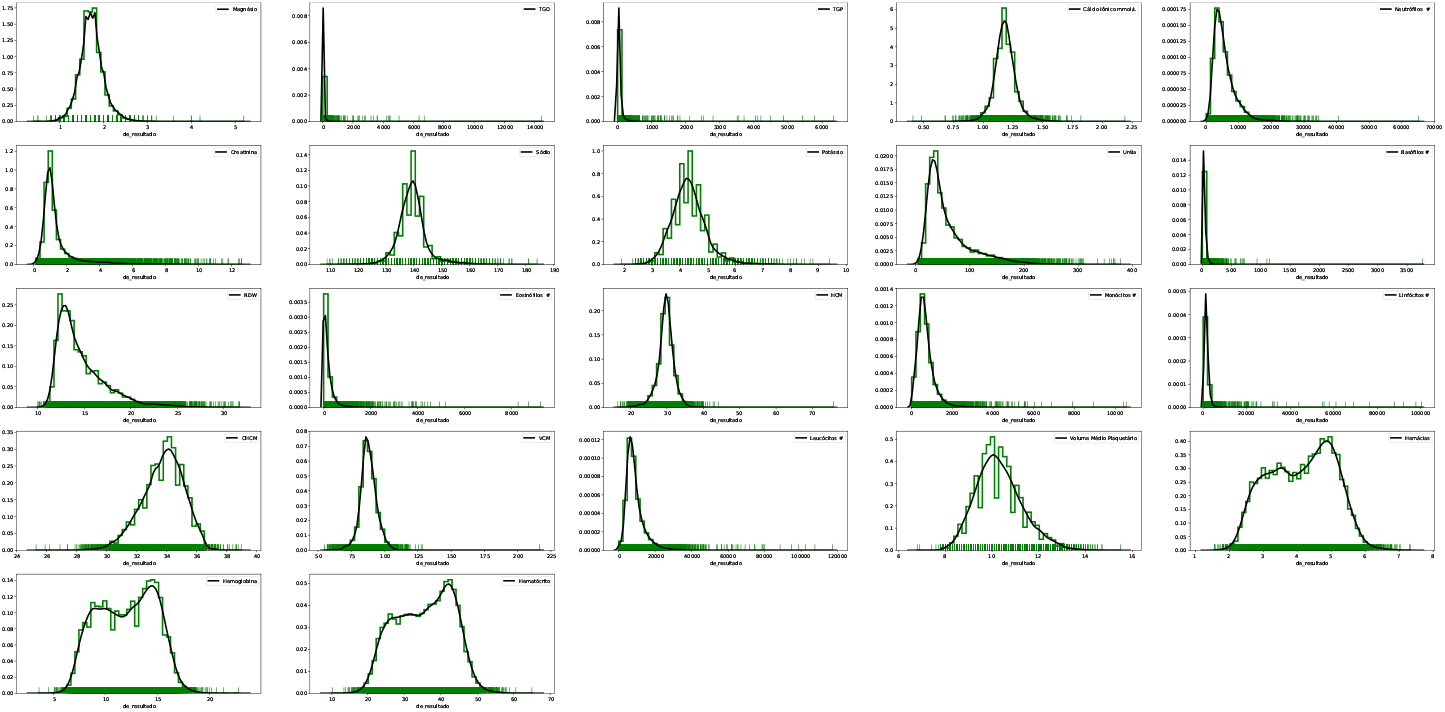
Men Analytes

The next graphic 13 present the distribution for positive cases of covid19. In the two previous images 12 and 13 is possible to observe a concentration of outliers in the sides of the normal distribution, i.e. TGO, TGP, Creatinina, Neutrofilos #, Ureia.

**Fig. 13.**
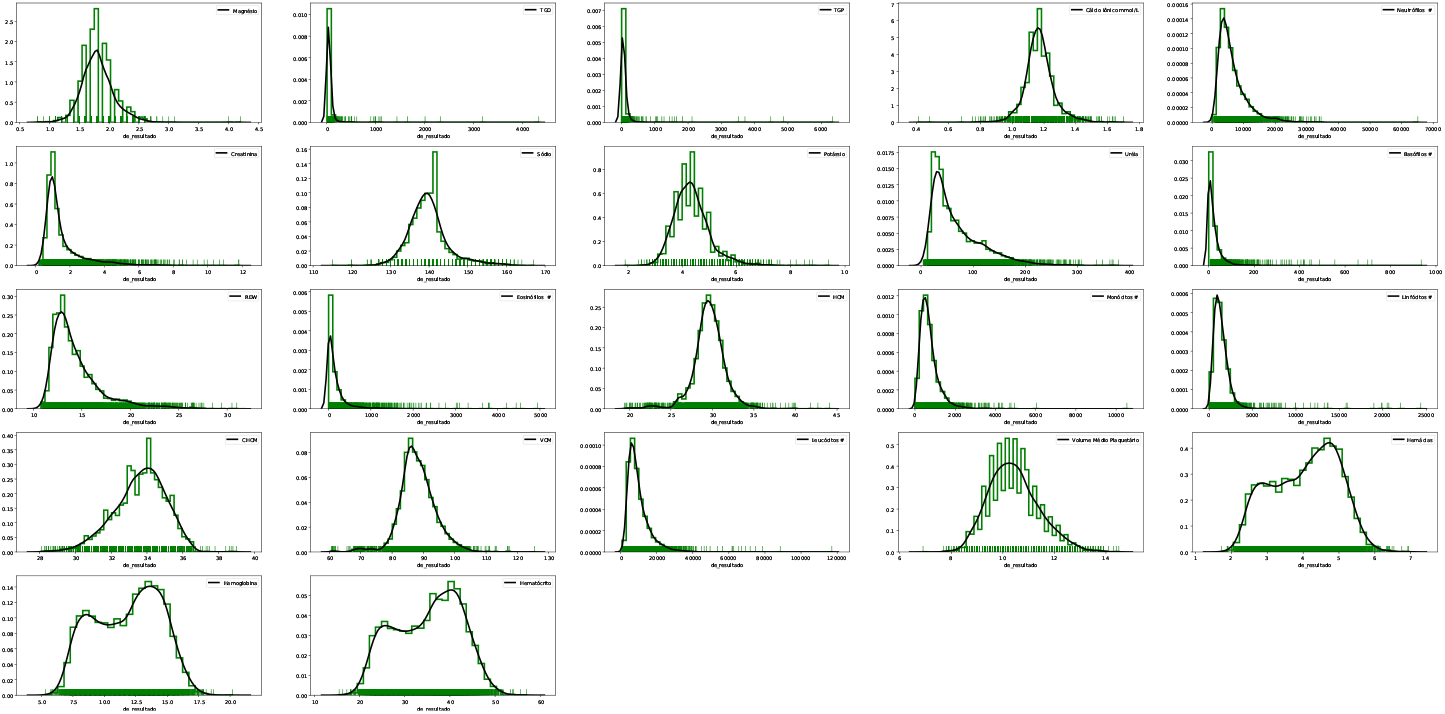
Men Analytes - Positives covid19 cases

And graphic 14 introduces the result after of cleaning values and considering patients with positive cases and the date when it was detected until it finishes or open(no date for discard test). Because the aim of the analysis is understand how is the behaviour of the patients with positive diagnosis of covid19 during the active phase of virus, from the start until the end. Analyzing, Fig. 14, it is possible to notice that the presence of outliers has disappeared, an exception with Basófilos #.

**Fig. 14.**
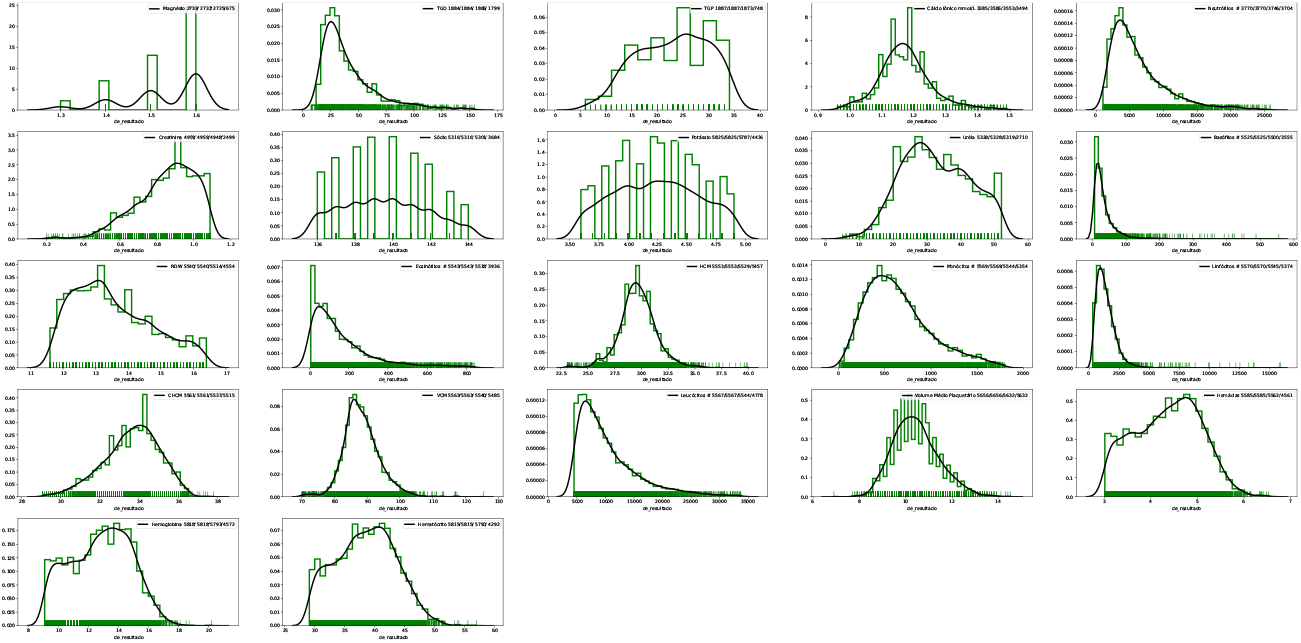
Filtered Men Analytes - Positives covid19 cases

Finally, Table 3.10 presentss the steps used to clean data and generate Fig. 14. First, only numerical values are considered, null values are discarded, and values out of reference range are not considered. For checking if values are inside of reference range, it was manually because there was many reference values too, only the lowest and highest value were used to filter data. Then, the reduction can be from 0.83 to 75.30 %. An initial number of exams was 108,152 and final value after filtering 86,814 with a reduction of almost 20% of the available data. Now, dataset is ready to answer more question and the research can continue.

**Table 6.** Reduction of Dataset

| de-analito | Initial | Only Numericals | Not null | Range | Reduction |
| --- | --- | --- | --- | --- | --- |
| Magnésio | 2733 | 2733 | 2725 | 675 | 75.30 |
| TGO | 1884 | 1884 | 1865 | 1799 | 4.51 |
| TGP | 1887 | 1887 | 1873 | 748 | 60.36 |
| Cálcio Iônico mmol/L | 3585 | 3585 | 3553 | 3494 | 2.54 |
| Neutrófilos # | 3770 | 3770 | 3746 | 3704 | 1.75 |
| Creatinina | 4959 | 4959 | 4948 | 2499 | 49.61 |
| Sódio | 5316 | 5316 | 5301 | 3684 | 30.70 |
| Potássio | 5825 | 5825 | 5787 | 4436 | 23.85 |
| Uréia | 5328 | 5328 | 5319 | 2710 | 49.14 |
| Basófilos # | 5525 | 5525 | 5500 | 3555 | 35.66 |
| RDW | 5540 | 5540 | 5514 | 4554 | 17.80 |
| Eosinófilos # | 5543 | 5543 | 5518 | 3936 | 28.99 |
| HCM | 5553 | 5553 | 5529 | 5457 | 1.73 |
| Monócitos # | 5569 | 5569 | 5544 | 5354 | 3.86 |
| Linfócitos # | 5570 | 5570 | 5545 | 5374 | 3.52 |
| CHCM | 5561 | 5561 | 5537 | 5515 | 0.83 |
| VCM | 5563 | 5563 | 5540 | 5485 | 1.40 |
| Leucócitos # | 5567 | 5567 | 5544 | 4778 | 14.17 |
| Volume Médio Plaquetário | 5656 | 5656 | 5632 | 5632 | 0.42 |
| Hemácias | 5585 | 5585 | 5562 | 4561 | 18.33 |
| Hemoglobina | 5818 | 5818 | 5793 | 4572 | 21.42 |
| Hematócrito | 5815 | 5815 | 5790 | 4292 | 26.19 |
| **Total** | **108152** |  |  | **86814** | **19.73** |

## 4 Conclusions

Coronavirus pandemic is active in the world, scientist are working to understand how to stop the virus, many areas are studying the covid19 impact in Heath, Economy therefore datasets related to patients are useful and important. Fapesp initiative to gather university and hospital is remarkable because it can foster research on the topic.

Real world datasets are not clean or ready for Data Mining or Data Science tasks then an exploratory phase is mandatory to see if data can be representative or useful to answer questions. Then, many cleaning steps were necessary to generate the final dataset and graphic, besides this cleaning step reduced the available dataset of men in 20%, with a maximum value of 75.30% for Magnesium Analyte, then it is possible a meanignful reduction of data is a cleaning task is performed.

Finally, share the process of analysis is useful for researchers interested to analyze with this dataset, so it can save time, effort to future research.

## 5 Recommendations

For researchers interested to work with these datasets, consider:

– Check if range of dates for each dataset to know if this data is useful for your study.
– Sirio-Libanes Hospital has some issues related to encoding, this is the smallest dataset then you must analyze if it useful for analysis and search for the problems to fix them.
– Only Einsteing dataset has a standardized output for covid19 exams: detected or not detected. If you are from Computer Science or related field, this is better for your study. Because, Fleury has a variety of outputs, therefore is necessary the presence or advice of one person related to Medicine to explain you the different values.
– If you want to automatize filtering considering reference range of values, remember there are many for many analytes, then the suggestion is check this manually to check if it is possible to code the process.

## 6 Future Work

For further work, a crossing of data is proposed to improve the analysis considering other variables, i.e. social-economic data, previous existence of health issues related to patients, considering data of other hospital to enhance the study. By the other hand, a deep analysis will be performed with this new cleaned dataset.

## Data Availability

The data used for this study is available on the site of author of the dataset: https://repositoriodatasharingfapesp.uspdigital.usp.br/

## Acknowledgement

The author wants to thank to Fabio Faria, professor of UNIFESP(Federal University of São Paulo) for the invitation to analyze this dataset, to the team DS-Covid for the discussion about the generated graphics during the data analysis task, more news about future will be available in: https://dscovid.github.io/

## Footnotes

1 Data extracted from website: https://virusncov.com/

